# Large Language Models Improve Coding Accuracy and Reimbursement in a Neonatal Intensive Care Unit

**DOI:** 10.1101/2025.07.23.25332086

**Authors:** Emma Holmes, Caroline Massarelli, Felix Richter, Stephanie Bernard, Robert Freeman, Nicholas Gavin, Courtney Juliano, Bruce D. Gelb, Benjamin S Glicksberg, Girish N Nadkarni, Eyal Klang

## Abstract

**Importance:** Diagnosis coding is essential for clinical care, research validity, and hospital reimbursement. In neonatal settings, manual coding is frequently error-prone, contributing to misclassification and financial losses. Large language models (LLMs) offer a scalable approach to improve diagnostic consistency and optimize revenue.

**Objective:** To compare the diagnostic accuracy of LLMs with human coders in identifying common neonatal diagnoses and assess the potential impact on revenue from Diagnosis-Related Group (DRG) assignment.

**Design:** This was a retrospective cross-sectional study conducted using data from 2022 to 2023. LLMs were prompted with all physician notes from the admission. Two neonatologists independently and blindly adjudicated diagnoses from three sources: human coders, GPT-4o, and GPT-o3-mini.

**Setting:** A single academic referral center’s neonatal intensive care unit (NICU).

**Participants:** The study included a consecutive sample of 100 infants admitted to the NICU who did not require respiratory support. All available physician notes from the hospital stay were included.

**Exposure:** Two HIPAA-compliant LLMs (GPT-4o and GPT-o3-mini) were prompted to assign diagnoses from a standardized list based on physician notes. Three prompt iterations were developed and reviewed for optimization prior to final evaluation.

**Main Outcomes and Measures:** The primary outcome was diagnostic accuracy compared with physician adjudication. Secondary outcomes included changes in expected DRG assignment and projected annual revenue.

**Results:** Among 100 infants (median gestational age 35.6 weeks, 52% male), GPT-o3-mini achieved 79.1% diagnostic accuracy (95% CI, 74.0%-84.2%), comparable to human coders at 76.3% (95% CI, 70.9%-81.7%; P = .52). GPT-4o underperformed at 58.6% (95% CI, 52.5%-64.7%; P < .001 vs both). Accuracy of GPT-o3-mini did not differ by DRG impact. Extrapolated to one year, correct GPT-o3-mini diagnoses yielded projected revenue of $5.71 million, compared to $4.82 million from human coders, an 18% increase.

**Conclusions and Relevance:** A HIPAA-compliant LLM demonstrated diagnostic accuracy comparable to human coders in neonatal billing while identifying higher-acuity diagnoses that improved projected reimbursement. LLMs may serve as effective adjuncts to manual coding in neonatal care, with potential clinical and financial benefit.

**Key Points:** *Question:* Can a large language model support accurate diagnosis generation for neonatal billing?

*Findings:* In this retrospective study of 100 neonates hospitalized in the Neonatal Intensive Care Unit, GPT-o3-mini demonstrated diagnostic accuracy comparable to human coders, as confirmed by physician review. Its implementation could yield an estimated 18% increase in revenue.

*Meaning:* Large language models may serve as effective adjuncts in neonatal coding, offering both diagnostic precision and financial benefit.

## 1. Introduction

Accurate medical coding is fundamental to modern healthcare systems. The International Classification of Diseases, 10th Revision (ICD-10), serves as the standard system for recording diagnoses and procedures across clinical settings. These codes inform billing and reimbursement and play an important role in operations, epidemiologic surveillance, and clinical research.^1,2^ Unfortunately, provider-documented ICD-10 codes are frequently inaccurate, which can undermine clinical decision support, impede patient care, introduce bias, and limit revenue capture.^3–6^

Neonatology is a field at particular risk for inaccuracy in diagnosis coding.^7^ Though research into the extent of the issue is limited, it has been shown that two of the most researched complications that occur in the neonatal intensive care unit (NICU), bronchopulmonary dysplasia and necrotizing enterocolitis, are not accurately coded.^8,9^ This is attributed to a combination of lack of granularity of neonatal diagnosis codes, resulting in overuse of broad codes, and inconsistent clinical definitions available in this relatively young and evolving field.^8,9^ Nonetheless ICD-10 codes continue to be routinely utilized to identify patient cohorts and, while they have utility, there is need for improved methods, particularly when studying rare diseases and using small samples.

In most U.S. healthcare systems, coding is generated through use of a combination of professional coders and computer-assisted coding (CAC) tools. Historically, CAC has relied on rule-based natural language processing (NLP) systems to scan the electronic health record and suggest diagnoses for review.^10^ Recent advances in large language models (LLMs) offer the potential to dramatically improve upon these tools. However, the adoption of LLMs for medical coding must proceed cautiously.^11^ Without rigorous evaluation, their use could replicate or even amplify existing inaccuracies, especially in sensitive domains like neonatology. To date there have not been any published studies examining the performance of LLMs in this field. In this study, we assessed the performance of LLMs in assigning ICD-10 codes for neonatal diagnoses and tested the hypothesis that LLMs would match or exceed the diagnostic accuracy of traditional manual coding in this setting.

## 2. Methods

### 2.1 Study Population and Data Sources

To maximize opportunities for diagnostic improvement, we targeted a population identified by a committee of neonatal physicians as being at elevated risk for under-diagnosis: infants who did not require respiratory support at any point during their hospitalization. We selected 100 consecutive admissions from the population of infants admitted to one level IV NICU within an academic health system in New York City between 2022 and 2023. ICD-10 codes, physician notes, and DRG were extracted from the electronic health record (EHR) for each patient. A list of relevant diagnoses was derived from the Centers for Medicare & Medicaid Services (CMS) DRG Definitions Manual by a committee of neonatal physicians and sorted into major and minor complications (**eTable 1 in Supplement**). This study was approved by the Mount Sinai Institutional Review Board under parent protocol 20-00338.

### 2.2 LLM Diagnosis Generation

We utilized Health Insurance Portability and Accountability Act-(HIPAA-) compliant GPT-4o and GPT-o3-mini via MSH’s secure Microsoft Azure tenant. We took a two-step approach to identifying relevant diagnoses with the LLMs: First, the models were prompted to examine each note for evidence of prematurity, low birth weight (LBW), and the common major complications; then the same note was re-examined for evidence of the minor complications. All unique diagnoses identified from the collection of patient notes were compiled (Figure 1). The initial prompt was provided to GPT-4o. We then underwent three iterations of prompting a small subset of patients to arrive at the ultimate prompt for GPT-o3-mini (**Appendix 1 in Supplement**). In all three cases, the performance of GPT-4o was worse, so data from the initial prompting of GPT-4o was utilized. We used default hyperparameter values for both models.

**Figure 1:**
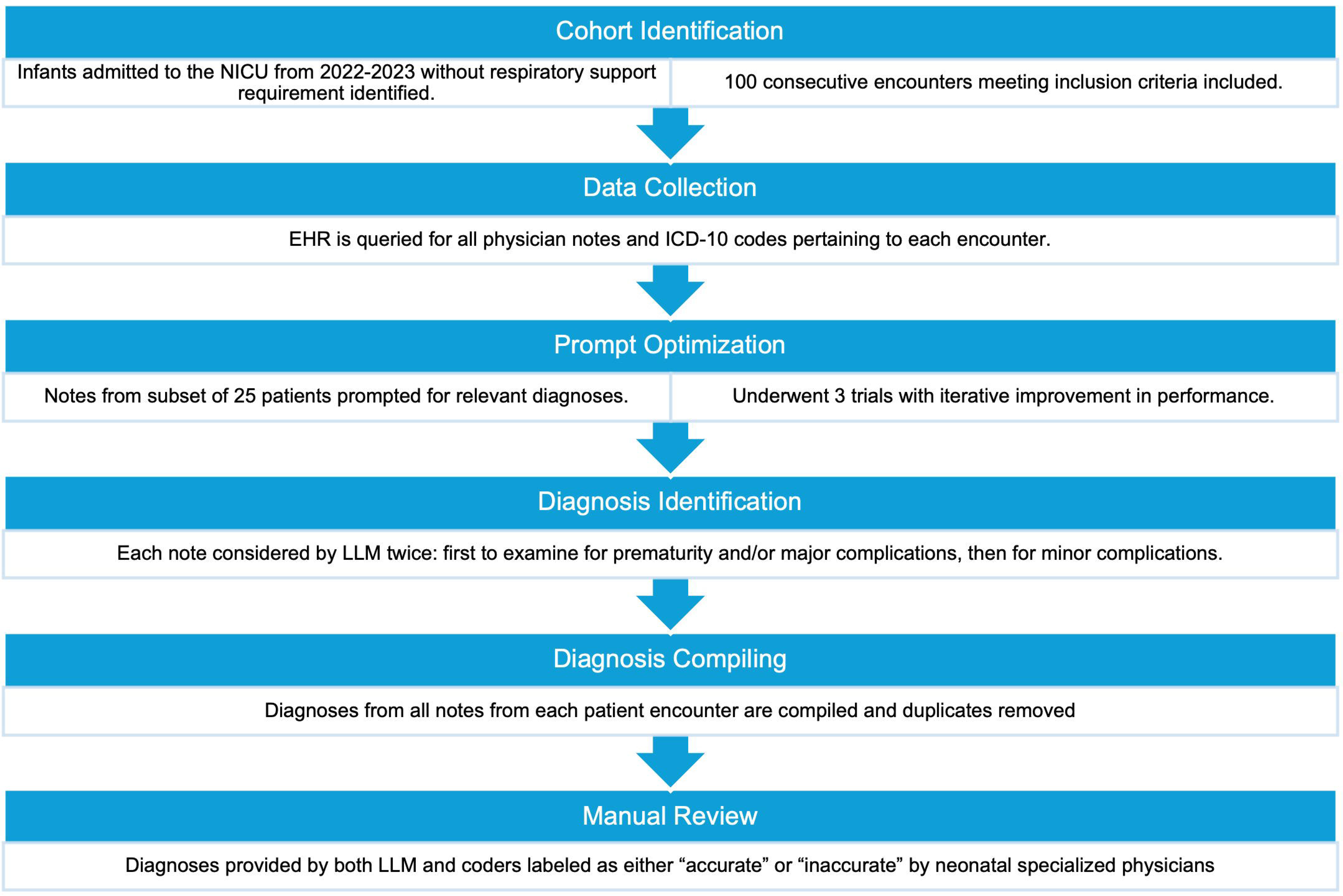
Methodology workflow diagram

### 2.3 Performance Evaluation

The compiled diagnoses from the three sources (Coders, GPT-4o, GPT-o3-mini) were each blindly labeled by two of three expert reviewers (CM, SB, and FR) as accurate or inaccurate. In the event of discordant labels, consensus was reached by the relevant pair. The expected impact on Diagnosis Related Group (DRG) was determined based on the CMS Definition Manual. Revenue estimation was based on Optum published national average DRG National Average Payment Table.^12^ Given that this is likely to be a tool that works alongside manual coders, rather than a replacement, we analyzed the revenue impact of only several iterations, including when only accurate codes are included.

### 2.4 Model Sensitivity Analysis

Additional analysis was performed on a separate subset of 50 patients, meeting the same inclusion criteria. This smaller analysis consisted of prompting GPT-4o only with a subset of the diagnoses that were ultimately used for the prompting of the larger cohort (**eTable 1 in Supplement**). All the diagnoses considered by the GPT-4o were labeled by the reviewers, regardless of whether the LLM detected them. The accuracy, sensitivity, specificity, positive predictive value (PPV), and negative predictive value (NPV) were then calculated from this cohort.

### 2.5 Projected Revenue and Cost Analysis

Projected revenue was estimated using the data from the publicly available National Average Payment table provided by Optum.^12^ LLM usage costs were estimated based on the assumption that each model reviewed 10 clinical notes per patient, which is conservative relative to the actual volume observed in this cohort. Each note was assumed to require approximately 2,000 input tokens and 20 output tokens per prompt, resulting in a total of 4,000,000 input tokens and 40,000 output tokens to run two prompts on all notes from 100 patient encounters. GPT-o3-mini also requires “reasoning tokens” which were estimated to be equivalent to the input tokens. Pricing was based on OpenAI’s published rates as of June 2025.^13^

### 2.6 Statistical Analysis

We reported accuracy for each diagnosis source and with subgroup analyses for impact on DRG and by individual diagnoses. Confidence intervals and p-values were obtained by pairwise z-tests. Inter-rater reliability was evaluated with pairwise Cohen’s Kappa coefficient. All analyses were performed in Python with open-source packages: pandas, sci-kit learn, and statsmodels. The models’ application programming interface (API) calls were done using the OpenAI package.

## 3. Results

### 3.1 Cohort Characteristics and Dataset

A cohort of 100 NICU patients was analyzed, of whom 62% were male. The median gestational age was 36.9 weeks (interquartile range [IQR], 4.0 weeks), and the median birth weight was 2.211 kg (IQR, 1.048 kg). The median length of stay was 6.5 days (IQR, 8.0 days). A total of 833 clinical notes were extracted from the electronic health records, yielding 2,510 unique diagnoses.

### 3.2 Comparative Performance

The accuracy for each source of diagnoses is shown in **Table 1**, along with a breakdown of accuracy by how the DRG is impacted. GPT-4o had statistically significantly worse accuracy than either of the other sources. GPT-o3-mini matched the performance of the coders (p = 0.52). There is no decrease in accuracy observed as the impact on DRG, and thus the expected revenue, increased.

**Table 1.** Accuracy and confidence intervals of diagnosis sources based on manual physician review.

| Source | Accuracy<br>[Confidence<br>Interval] | Accuracy [CI] by DRG Impact |  |  |
| --- | --- | --- | --- | --- |
|  |  | Improves | Agrees | Decreases |
| **Coders** | 0.763<sup>a</sup><br>[0.735-0.788] |  |  |  |
| **GPT-4o** | 0.586<br>[0.551-0.620] | 0.653<br>[0.597-0.705] | 0.569<sup>b</sup><br>[0.519-0.618] | 0.512<sup>b</sup><br>[0.368-0.654] |
| **GPT-o3-mini** | 0.791<sup>a</sup><br>[0.761-0.819] | 0.797<br>[0.746-0.839] | 0.809<br>[0.766-0.845] | 0.703<br>[0.542-0.825] |

Threshold for statistical significance $p < 0.05$ .
<sup>a</sup> Statistically significant difference between indicated model and GPT-4o
<sup>b</sup> Statistically significant difference between indicated accuracy and accuracy for improvement in DRG

The count and accuracy of the diagnoses identified in at least 10 patient charts are displayed in **Table 2**. There is improvement overall in the performance of GPT-o3-mini in most diagnoses, but there remain several diagnoses with less than 50% accuracy (**Table 2**).

**Table 2.** Counts and accuracy for each model stratified by diagnosis, limited to the diagnoses identified in at least 10 patients by at least one of the LLMs.

| **Diagnosis** | **GPT-4o**<br>**Count** | **GPT-4o**<br>**Accuracy** | **GPT-o3-**<br>**mini-Count** | **GPT-o3-mini**<br>**Accuracy** |
| --- | --- | --- | --- | --- |
| Preterm newborn, unspecified weeks of gestation | 51 | 1.000 | 50 | 1.000 |
| Newborn (suspected to be) affected by Cesarean delivery | 47 | 0.979 | 47 | 0.979 |
| Other low birth weight newborn, unspecified weight | 44 | 0.932 | 40 | 1.000 |
| Disturbances of sodium balance of newborn | 44 | 0.727 | 34 | 0.882 |
| Disturbances of potassium balance of newborn | 37 | 0.243 | 27 | 0.222 |
| Syndrome of infant of a diabetic mother | 36 | 0.194 | 17 | 0.471 |
| Newborn affected by abnormality in fetal heart rate or rhythm during labor | 33 | 0.727 | 28 | 0.929 |
| Newborn affected by malpresentation before labor | 32 | 0.406 | 15 | 0.933 |
| Other respiratory distress of newborn | 29 | 0.241 | 6 | 0.500 |
| Other transitory electrolyte disturbances of newborn | 29 | 0.931 | 3 | 1.000 |
| Newborn affected by maternal hypertensive disorders | 28 | 0.821 | 26 | 0.923 |
| Transient tachypnea of newborn | 25 | 0.080 | 20 | 0.350 |
| Metabolic acidemia noted at birth | 25 | 0.400 | 11 | 0.909 |
| Other neonatal hypoglycemia | 23 | 0.565 | 53 | 0.868 |
| Newborn affected by maternal complication of pregnancy, unspecified | 23 | 0.522 | 34 | 0.882 |
| Newborn (suspected to be) affected by premature ROM | 22 | 0.364 | 15 | 0.533 |
| Other apnea of newborn | 22 | 0.545 | 3 | 1.000 |
| Newborn affected by slow intrauterine growth, unspecified | 21 | 0.714 | 23 | 0.826 |
| Disturbance of temperature regulation of newborn, unspecified | 20 | 1.000 | 56 | 1.000 |
| Newborn affected by other maternal medication | 20 | 0.600 | 10 | 0.900 |
| Neonatal bradycardia | 15 | 0.667 | 28 | 0.857 |
| Disturbance of cerebral status of newborn, unspecified | 14 | 0.500 | 15 | 0.267 |
| Newborn (suspected to be) affected by oligohydramnios | 14 | 0.286 | 4 | 1.000 |
| Late metabolic acidosis of newborn | 11 | 0.091 | 11 | 0.727 |
| Other neonatal hypocalcemia | 10 | 0.900 | 18 | 1.000 |
| Respiratory failure of newborn | 10 | 0.400 | 12 | 0.667 |
| Cyanotic attacks of newborn | 8 | 0.625 | 44 | 0.341 |
| Neonatal jaundice associated with preterm delivery | 6 | 0.167 | 25 | 0.760 |
| Newborn esophageal reflux | 5 | 0.600 | 11 | 1.000 |

### 3.3 Interrater Reliability

Cohen’s Kappa coefficients between Reviewer 1 and 2 was 0.427 and between Reviewer 1 and 3 was 0.203. Reviewers 2 and 3 never labeled the same diagnoses. These values are less than desirable for a gold standard, indicative of ambiguity in several of the diagnoses being considered.

### 3.4 Sensitivity Analysis

In our sensitivity analysis cohort of 50 similar patients, GPT-4o and coders had comparable accuracy (73.6% vs. 71.1%, respectively). GPT-4o achieved high specificity, ranging from 0.81-1, but variable sensitivity, ranging from 0-0.85, in the subset diagnoses included in the prompting (**eTable 2 in Supplement**). There was strong positive predictive value (PPV) in most cases, with notable exception of “Phototherapy treatment for hyperbilirubinemia” and “Leukocytosis.”

### 3.5 DRG and Revenue Impact

Expected DRG based on LLM diagnoses was shifted towards higher acuity for most patients, due primarily to rise in classification of admissions as 791 (Prematurity with Major Problems) and 793 (Full term Neonate with Major Problems) (**Figure 2**). Upon implementation this would result in a substantial increase in projected revenue, which is robust to exclusion of all diagnoses deemed inaccurate by the reviewers (**Table 3**). Extrapolating the outcomes to 1 year and including only accurate GPT-o3-mini diagnoses the expected revenue would be $5,712,254.46 compared to $4,824,574.76 based on the coder’s output, signifying a potential increase in revenue by 18%.

**Figure 2:**
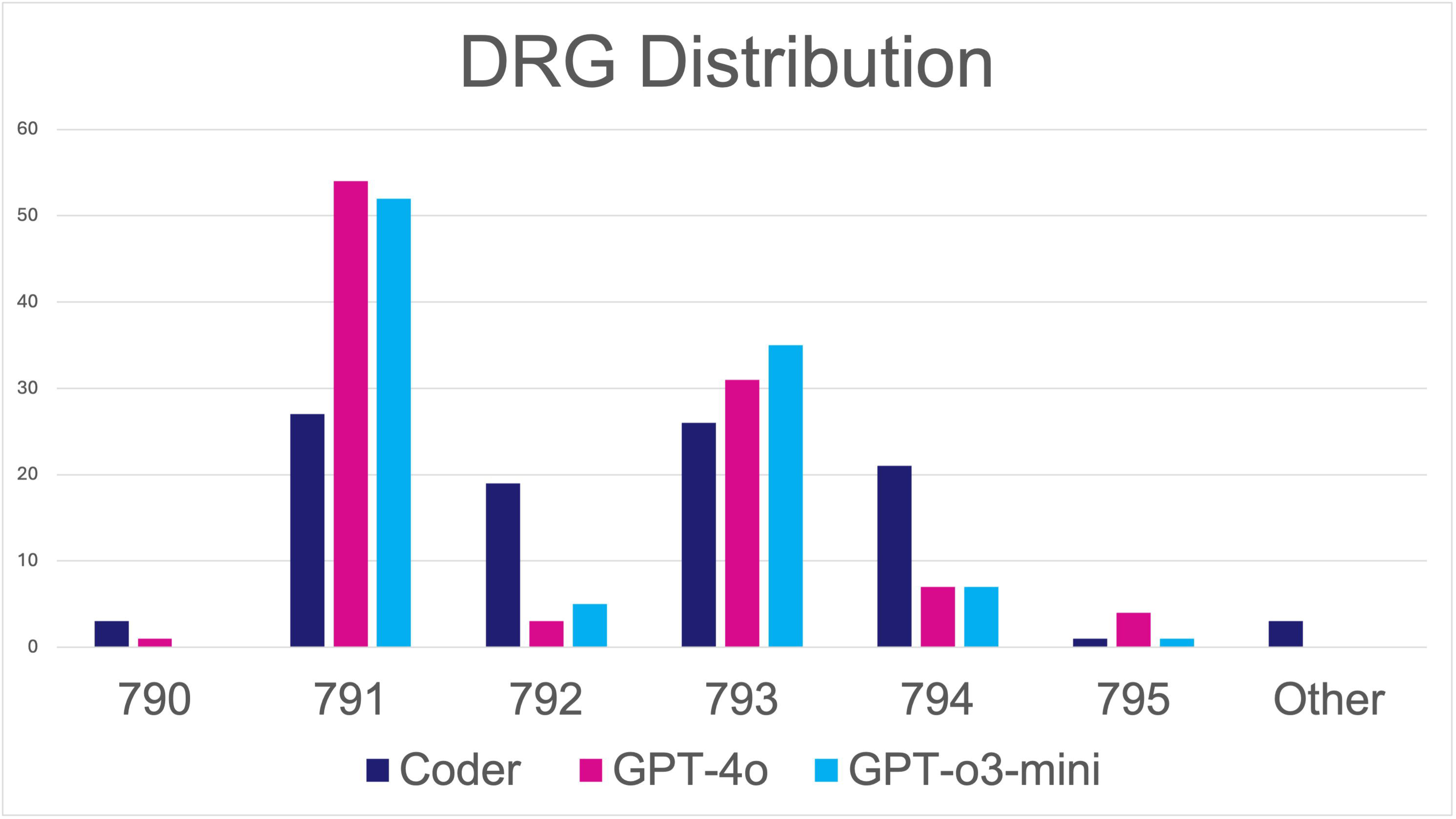
Distribution of expected DRG for each of the sources of diagnosis codes

**Table 3.** Expected revenue (gross and change) based on the diagnoses suggested by each source for this 100-baby sample.

| **Source** | **Gross Revenue** | **Increase from Base** |
| --- | --- | --- |
| **Coders** | \$1,754,390.82 | N/A |
| **GPT-4o** | \$2,095,731.51 | \$341,340.69 |
| **GPT-o3-mini** | \$2,134,579.73 | \$380,188.91 |
| **GPT-o3-mini (only accurate diagnoses)** | \$2,077,183.44 | \$322,792.62 |

### 3.6 Cost Analysis

The LLMs usage in this study incurred an estimated conservative cost of $10.40 and $6.78 for GPT-4 and GPT-o3-mini, respectively (**eTable 3 in Supplement**).

## 4. Discussion

Our study demonstrates that GPT-o3-mini can match the diagnostic accuracy of human coders in low-complexity NICU patients, while also shifting DRG profiles and increasing projected revenue. GPT-4o was less accurate than both GPT-o3-mini and coders when provided with the same prompting. The performance gap highlights the need for rigorous, modell1lspecific evaluation. Our study underlines the importance of rigorous testing prior to deployment of any such model for this purpose.

In this case, we suspect that the superior performance of GPT-o3-mini can be attributed to its improved reasoning capability.^14^ The task of medical diagnosis is complex: it requires temporal reasoning, detection of negative language, and causal inference based on clinical signs and symptoms. GPT-o models have been extensively trained on “chain of thought” rich datasets, which has been consistently shown to improve the LLM’s capability for logical consistency and structured thinking.^15,16^

Our results are supported by a body of research that has suggested sufficient or even superior diagnostic performance of LLMs when compared to physicians.^17–19^ These findings are influenced heavily by subspecialty, methodology for prompting and optimizing the LLM, as well as by the evaluation technique.^20^ A recent meta-analysis found that LLMs slightly underperform clinical specialists at diagnosis, but there is considerable variability in performance.^21^ This suggests that the differing methodologies may provide insight into optimization techniques, and the need for individualization of the approach based on the clinical task.

Multiple medical-coding specific studies have demonstrated worse performance by the LLM compared to coding specialists, which conflicts with our results.^22^ ^23^ Of note, these studies are performed among adult populations and, thus, have different baseline performance among coders and CAC software, which are historically trained on adult medicine. The work of Mustafa et al. further demonstrates the variability of performance of human coders; they identified individual medical coders that outperformed their LLM (GPT-4o), but when compared to the median coder performance, the LLM matched human accuracy.^23^ In contrast, our study aligns with the prior findings of our group in the adult emergency department, where a slightly different approach including retrieval-augmented generation was necessary to aid the LLM in achieving adequate performance.^24^ This again underscores the importance of targeted methodology in differing populations and clinical tasks.

The NICU population is an important target for improvement because of the unique challenge of diagnosis. The challenge is attributable both to the requirement of domain knowledge of relatively unique diagnoses and to the somewhat subjective definitions of the relevant diagnoses. The subjectivity of NICU diagnoses is underscored by the significant inter-rater variability observed between the expert reviewers in this study: despite years of training, there remain discrepancies in diagnostic consideration for even these more common NICU diagnoses. This subjectivity explains why the diagnosis was the largest determining factor of accuracy for the LLMs in our study. While this may lead some to believe that a human coder is appropriate for this complex and nuanced task, our results suggest that the specific guidance provided in the prompting, when coupled with a reasoning LLM model, was able to match the performance of these human coders. Prospective study with a larger cohort (to better characterize the rarer conditions) would clarify the most appropriate role of LLMs in this space.

Employment of LLMs may be limited in clinical settings due to lack of transparency or trust in the “black box” models. Multiple approaches can be taken to address this, such as asking the LLM to provide the text from the medical record that supports their diagnosis or requirement of multiple instances of supportive evidence to avoid false positives. There are also implementation barriers to inserting the LLM-derived code into the provider workflow. A key principle is keeping the “human in the loop.” Rather than considering these models as autonomous agents, they should be considered assistants, perhaps replacing the outdated CAC technologies that are already deployed in medical systems throughout the country.

Our study is limited by extrapolation of DRG and revenue from national sources and, thus, cannot be considered as an actual examination of the revenue impact of LLMs. In practice, these diagnoses would be reviewed by coding experts and then submitted to payors prior to having an impact on revenue. By design, this retrospective study merely suggests promise but cannot assess the true impact of implementing an LLM into NICU coding workflow. Moreover, our study is limited to a single center. Future research will entail a pilot program of the GPT-o3-mini model as a substitute for existing CAC technology. We did not employ techniques such as fine-tuning and retrieval augmented generation (RAG). LLM research is moving fast, and it is likely that newer models will perform even better than GPT-o3-mini.

## Data Availability

Data is not available due to privacy concerns

