## Supplement for "Large Language Models Improve Coding Accuracy and Reimbursement in a Neonatal Intensive Care Unit"

Supplemental Materials

### **Supplemental Table 1**

| **Major Complications** | **Minor Complications** |
| --- | --- |
| P034: Newborn (suspected to be) affected by Cesarean delivery | P000: Maternal hypertensive disorders |
| P392: Intra-amniotic infection | P011: Premature rupture of membranes |
| P704: Other neonatal hypoglycemia | P012: Oligohydramnios |
| P742: Disturbances of sodium balance | P013: Polyhydramnios |
| P743: Disturbances of potassium balance | P017: Malpresentation before labor |
| P744: Other transitory electrolyte disturbances | P019: Maternal complication of pregnancy, unspecified |
| P741: Dehydration of newborn | P03811: Abnormal fetal heart rate or rhythm |
| P711: Other neonatal hypocalcemia | P0382: Meconium passage during delivery |
| P712: Neonatal hypomagnesemia | P041: Affected by other maternal medication |
| P740: Late metabolic acidosis | P059: Slow intrauterine growth, unspecified |
| P919:  Disturbance of cerebral status | P192: Metabolic acidemia at birth |
| P252: Pneumomediastinum | P221: Transient tachypnea |
| P610: Transient neonatal thrombocytopenia | P228: Other respiratory distress |
| P280: Primary atelectasis | P284: Other apnea of newborn |
| P293: Persistent fetal circulation | P282: Cyanotic attacks |
| P369: Bacterial sepsis of newborn, unspecified | P289: Respiratory condition, unspecified |
| P559: Hemolytic disease of newborn, unspecified | P2911: Neonatal tachycardia |
| P508: Newborn affected by other intrauterine blood loss | P2912: Neonatal bradycardia |
| P251: Pneumothorax | P550: Rh isoimmunization |
| P285: Respiratory failure of newborn | P551: ABO isoimmunization |
|  | P590: Neonatal jaundice associated with preterm |
|  | P611: Polycythemia neonatorum |
|  | P615: Transient neonatal neutropenia |
|  | P701: Infant of a diabetic mother |
|  | P761: Transitory ileus |
|  | P7883: Newborn esophageal reflux |
|  | P819: Disturbance of temperature regulation, unspecified |

**eTable1:** List of complication diagnoses being considered by LLMs, stratified by severity

### **Appendix 1**

**Complete prompting strategy:**

def func_prompt_1 (text):

    prompt = f"""

Hello GPT-4, you are given a NICU provider note. Please identify which of the following diagnoses are applicable to the patient, based on the criteria provided.

If a condition is met, return its code and a supporting sentence from the text.

Return all findings as a single JSON array. If none are found, return "[]".

Conditions with codes and criteria to be considered:

Low birth weight newborn under 2.5 kg: P0710. To qualify for this diagnosis, the infant must have a birth weight below 2.5 kg

Preterm newborn under 37 weeks: P0730. To qualify for this diagnosis, the infant must have been born at gestational age less than 37 weeks

Cesarean delivery: P034. To qualify for this diagnosis, the infant must have been born via cesarean section

Intra-amniotic infection: P392. To qualify for this diagnosis, there must be mention of maternal chorioamnionitis

Other neonatal hypoglycemia: P704. To qualify for this diagnosis, the infant must have had at least 2 blood sugar checks, with at least 1 falling below 50

Disturbances of sodium balance: P742. To qualify for this diagnosis, there must be at least 2 sodium (Na) levels checked, with at least one of the levels being outside the normal range (134-145). Please disregard values from blood gases (i.e. NAART, NAVEN, NACAP).

Disturbances of potassium balance: P743. To qualify for this diagnosis, there must be at least 2 potassium (K) levels checked, with at least one of the levels being outside the normal range (3-6.6).

Other transitory electrolyte disturbances: P744. To qualify for this diagnosis, there must be at least 2 checks of a given electrolyte, with at least one of the levels being outside the normal range for that electrolyte. Please exclude elevated chloride levels.

Dehydration of newborn: P741. To qualify for this diagnosis, the infant must be noted to require supplemental fluids, which can be with formula or IV fluids. Please don’t use this diagnosis if the IV fluids are started for hypoglycemia.

Other neonatal hypocalcemia: P711. To qualify for this diagnosis, there must be at least 2 checks calcium (Ca or iCal), with at least one of the levels being below the lower limit of normal (8 for non-ionized, 1 for ionized)

Neonatal hypomagnesemia: P712. To qualify for this diagnosis, there must be at least 2 checks magnesium, with at least one of the levels being below the lower limit of normal (1.5)

Late metabolic acidosis: P740. To qualify for this diagnosis, the infant must have evidence of metabolic acidosis (either a bicarbonate level less than 18 or a base excess of less than –5) that occurs after 3 days of life with a subsequent repeat that is improved

Disturbance of cerebral status: P919. To qualify for this diagnosis, the infant must be noted to have a period of abnormal neurologic status, which could be abnormal eye, limb, or whole-body movements, lip smacking, decreased responsiveness, or another indication of abnormal neurologic status

Pneumomediastinum: P252. To qualify for this diagnosis, the newborn must have an x-ray in which pneumomediastinum is present

Transient neonatal thrombocytopenia: P610. To qualify for this diagnosis, the newborn must have a low value for platelets (that is less than 150), which is checked at least once more and noted to improve

Primary atelectasis: P280. To qualify for this diagnosis, the newborn must have an x-ray in which atelectasis is present

Persistent fetal circulation: P293. To qualify for this diagnosis, there must be concern for pulmonary hypertension documented

Bacterial sepsis of newborn, unspecified: P369. To qualify for this diagnosis, the infant must have a positive blood culture

Hemolytic disease of newborn, unspecified: P559. To qualify for this diagnosis, the infant must have evidence of hemolysis, such as dropping hematocrit and rising bilirubin

Newborn affected by other intrauterine blood loss: P508. To qualify for this diagnosis, the infant must have had a hematocrit below 35 on the first check after birth

Pneumothorax: P251. To qualify for this diagnosis, the newborn must have an x-ray in which a pneumothorax is present

Respiratory failure of newborn: P285. To qualify for this diagnosis, the infant must require respiratory support, such as CPAP, during their hospitalization

For example:

[{{"code":"P034", "sentence":"Infant placed under radiant warmer."}},

{{"code":"P251", "sentence":...]

\#### Note:

{text}

"""

    return prompt

def func_prompt_2 (text):

    prompt = f"""

Hello GPT-4, you are given a NICU provider note. Please identify which of the following diagnoses are applicable to the patient, based on the criteria provided.

If a condition is met, return its code and a supporting sentence from the text.

Return all findings as a single JSON array.

If none are found, return "[]".

Conditions with codes and criteria to be considered:

Maternal hypertensive disorders: P000. To qualify for this diagnosis, the mother of the newborn must have been noted to have pre-eclampsia or other hypertensive disorder

Premature rupture of membranes: P011. To qualify for this diagnosis, the mother of the newborn must have been noted to have rupture of membranes prior to the onset of labor

Oligohydramnios: P012. To qualify for this diagnosis, the mother of the newborn must have been noted to have oligohydramnios

Polyhydramnios: P013. To qualify for this diagnosis, the mother of the newborn must have been noted to have polyhydramnios

Malpresentation before labor: P017. To qualify for this diagnosis, the baby must have been in breech or other non-vertex position

Maternal complication of pregnancy, unspecified: P019

Abnormal fetal heart rate or rhythm: P03811. To qualify for this diagnosis, there must have been category 2 fetal heart rate or non-reassuring fetal heart rate during labor (aka C2FHR, C2FHT, cat 2, NRFHT)

Meconium passage during delivery: P0382. To qualify for this diagnosis, there must have been meconium stained amniotic fluid

Affected by other maternal medication: P041. To qualify for this diagnosis, the mother of the infant must have been taking a medication known to affect fetuses (such as SSRIs, benzodiazepines, opiates, anti-epileptic medications)

Slow intrauterine growth, unspecified: P059. To qualify for this diagnosis, there must be documentation that mentions intra-uterine growth restriction or IUGR

Metabolic acidemia at birth: P192. To qualify for this diagnosis, the infant must have either cord gases or initial blood gas after birth with base excess less than -8 and a repeat blood gas must be obtained

Transient tachypnea: P221. To qualify for this diagnosis, the infant must have been admitted to the NICU for concern for respiratory distress, which can be characterized by grunting, tachypnea, or retractions, that resolved rapidly

Other respiratory distress: P228. To qualify for this diagnosis, the infant must have been noted to have respiratory distress, but is not noted to have: transient tachypnea of the newborn, respiratory distress syndrome, or pneumothorax

Cyanotic attacks: P282. To qualify for this diagnosis, the infant must have been noted to have desaturations or cyanotic episodes

Other apnea of newborn: P284. To qualify for this diagnosis, the infant must have been born full term and have episodes concerning for apnea

Respiratory condition, unspecified: P289. To qualify for this diagnosis, the infant must have had concerning respiratory exam findings, but no clear diagnosis made that would qualify the patient for a different diagnosis

Neonatal tachycardia: P2911. To qualify for this diagnosis, there must be comment regarding tachycardia in the documentation

Neonatal bradycardia: P2912. To qualify for this diagnosis, there must be comment regarding bradycardia in the documentation

Rh isoimmunization: P550. To qualify for this diagnosis, the infant must be coombs+ and have a different Rh group than their mother, that is mother Rh negative and the infant is Rh positive

ABO isoimmunization: P551. To qualify for this diagnosis, the infant must be coombs+ and have a different blood type than their mother, that is mother is O blood type and infant is either A, B, or AB blood type

Neonatal jaundice associated with preterm: P590. To qualify for this diagnosis, the infant must require phototherapy for hyperbilirubinemia and also have been born before gestational age 37 weeks

Polycythemia neonatorum: P611. To qualify for this diagnosis, the infant must have a hematocrit greater than 65

Transient neonatal neutropenia: P615. To qualify for this diagnosis, the infant must have had one instance of an absolute neutrophil count less than 1500, which improved

Infant of a diabetic mother: P701. To qualify for this diagnosis, the mother of the infant must have had record of being diabetic and the infant must have some manifestation of that condition, which could be: respiratory distress, cardiomegaly, hypoglycemia, hypocalcemia, or other congenital anomalies

Transitory ileus: P761. To qualify for this diagnosis, there must be concern for delayed passage of meconium, constipation, or ileus documented

Newborn esophageal reflux: P7883. To qualify for this diagnosis, the infant must have multiple episodes of emesis documented

Disturbance of temperature regulation, unspecified: P819. To qualify for this diagnosis, the infant must have had one temperature recorded outside the normal range (36.5-37.9)

For example:

[{{"code":"P221", "sentence":"Noted mild respiratory difficulty without intervention."}},

{{"code":"P7883", "sentence":... ]

\#### Note:

{text}

"""

    return prompt

### **Supplemental Table 2**

| Diagnosis | Sensitivity | Specificity | PPV | NPV |
| --- | --- | --- | --- | --- |
| ‘ABO incompatibility’ | 0.78 | 0.98 | 0.88 | 0.95 |
| ‘Neonatal hypoglycemia’ | 0.76 | 0.92 | 0.90 | 0.79 |
| ‘Slow feeding newborn’ | 0.85 | 0.81 | 0.61 | 0.94 |
| ‘GE reflux, neonatal’ | 0.05 | 1 | 1 | 0.63 |
| ‘Temperature instability in newborn’ | 0.59 | 0.88 | 0.91 | 0.50 |
| ‘Phototherapy treatment for hyperbilirubinemia’ | 0 | 1 | N/A | 0.84 |
| ‘Hyponatremia of newborn’ | 0.22 | 1 | 1 | 0.85 |
| ‘Leukocytosis’ | 1 | 0.82 | 0.1 | 1 |
| ‘Need for observation and evaluation of newborn for sepsis’ | 0.18 | 0.96 | 0.8 | 0.6 |
| ‘Metabolic acidosis in newborn’ | 0.29 | 1 | 1 | 0.90 |

**eTable2.** Sensitivity of GPT-4o on cohort of 50 NICU patients for subset of relevant diagnoses. PPV = positive predictive value, NPV = negative predictive value.

### **Supplemental Table 3**

|  | **GPT-4o** | **GPT-o3-mini** |
| --- | --- | --- |
| **Input** | $2.50/1M tokens | $1.10/1M tokens |
| **Reasoning** | N/A | $0.55/1M tokens |
| **Output** | $10.00/1M tokens | $4.40/1M tokens |
| **Estimated total cost (per 100 patients)** | $10.40 | $6.78 |

**eTable3**. Cost of LLM usage for 100 patients, assuming 10 notes per patient, 1,000 input tokens per prompt, 20 output tokens per prompt, and 2 prompts per note, resulting in a total of 4M input tokens, 40,000 output tokens, and 4M reasoning tokens (applicable to GPT-o3-mini only).
